# Genetic Architecture of Postpartum Psychosis: From Common to Rare Genetic Variation

**DOI:** 10.1101/2024.12.09.24318732

**Authors:** Seulgi Jung, Madison Caballero, Adrianna Kępińska, Shelby Smout, Trine Munk-Olsen, Thalia K. Robakis, Veerle Bergink, Behrang Mahjani

**Affiliations:** Seaver Autism Center for Research and Treatment, Icahn School of Medicine at Mount Sinai, New York, NY, USA; Department of Psychiatry, Icahn School of Medicine at Mount Sinai, New York, NY, USA; Department of Genetics and Genomic Sciences, Icahn School of Medicine at Mount Sinai, New York, NY, USA; Department of Clinical Research, Research Unit Children and Adolescent Psychiatry, University of Southern Denmark, Denmark; Department of Psychiatry, Erasmus Medical Center, Rotterdam, The Netherlands; Department of Artificial Intelligence and Human Health, Icahn School of Medicine at Mount Sinai, New York, NY, USA; Mindich Child Health and Development Institute, Icahn School of Medicine at Mount Sinai, New York, NY, USA; Department of Molecular Medicine and Surgery, Karolinska Institutet, Stockholm; Department of Medical Epidemiology and Biostatistics, Karolinska Institutet, Stockholm, Sweden

**Keywords:** Postpartum psychosis, Genetic basis, Common genetic variation, Rare genetic variants, Family-based heritability

## Abstract

Postpartum psychosis is a severe psychiatric condition marked by the abrupt onset of psychosis, mania, or psychotic depression following childbirth. Despite evidence for a strong genetic basis, the roles of common and rare genetic variation remain poorly understood. Leveraging data from Swedish national registers and genomic data from the All of Us Research Program, we estimated family-based heritability at 55% and WGS-based heritability at 37%, with an overrepresentation on the X chromosome. Rare coding variant analysis identified *DNMT1* and *HMGCR* as potential risk genes (q < 0.1). Analysis of 240,009 samples from All of Us demonstrated significant associations between these genes and multiple psychiatric disorders, supporting their biological relevance. Additionally, 17% of bipolar disorder, 21% of schizophrenia, and 16–25% of multiple autoimmune disorder risk genes overlapped with postpartum psychosis. These findings reveal unique genetic contributions and shared pathways, providing a foundation for understanding pathophysiology and advancing therapeutic strategies.

## Introduction

Postpartum psychosis is a rare yet profoundly severe psychiatric condition, affecting 0.1-0.2% of mothers following childbirth.^1,2^ It is marked by an acute onset, typically occurring within days to weeks postpartum, and is characterized by a spectrum of symptoms, including mania, depression, psychosis, cognitive disorganization, irritability, and sleep disturbances.^3^ These symptoms pose significant risks of suicide and infanticide, necessitating immediate medical intervention. This condition also highlights a striking vulnerability in maternal mental health, with a 10-fold increase in the rate of first-onset psychiatric hospitalizations during this period, encompassing unipolar depressive disorders, schizophrenia, and bipolar affective disorders.^4^ Although postpartum psychosis is classified as a bipolar spectrum disorder, its distinct timing, triggers, and associated risks underscore the need for a specialized classification and approach in both research and clinical practice.

The underlying pathophysiology of postpartum psychosis, particularly its genetic basis, remains poorly understood. Emerging evidence points to structural brain changes,^5^ neuroimmune dysregulation,^6,7^ and alterations in the hypothalamic-pituitary-adrenal axis as being associated with this disorder.^8^ Genetic contributions to the etiology of postpartum psychosis are becoming increasingly evident. The only genetic study to date used data from women in the Bipolar Disorder Research Network database consisted of 203 mothers experiencing their first episode of postpartum psychosis and 1,225 parous women with a history of bipolar disorder.^9^ This study revealed that women with first-onset postpartum psychosis had polygenic risk scores (PRSs) similar to those with bipolar disorder, both higher than controls. Yet, unlike the PRS for bipolar disorder, the PRS for postpartum psychosis did not overlap with that of major depression, suggesting postpartum psychosis may have a distinct genetic etiology. This study highlights the need for more postpartum psychosis-specific studies.

Building on this insight, our recent study leveraging family data from Swedish national registers, estimated a relative recurrence risk of 10.69 (95% confidence interval = 6.60 – 16.26) among sisters of mothers diagnosed with their first episode of postpartum psychosis.^10^ This rate is higher than the familial recurrence risk for bipolar disorder, which is documented at 7.9 (95% = 7.1 – 8.8) in similar Swedish registry studies.^11^ These findings further support the hypothesis that postpartum psychosis has a substantial genetic basis, overlapping with but distinct from other well-studied psychiatric disorders.

Despite growing evidence for genetic contributions to postpartum psychosis, its rarity poses significant challenges in assembling sufficiently large cohorts to identify specific causal variants. Common genetic variants, which contribute to polygenic risk for most psychiatric disorders, including postpartum psychosis, typically have low to modest effect sizes. Detecting associations with these variants requires very large sample sizes, which are often infeasible to achieve for rare disorders. In contrast, rare deleterious variants, such as loss-of-function or damaging missense mutations, often have larger effect sizes that facilitate their detection in smaller cohorts. Identifying these rare variants, however, requires robust statistical methods that account for their low frequency and functional relevance. Bayesian approaches, such as the Transmission and De Novo Association (TADA) test, address some of these challenges by integrating prior biological knowledge, pathogenicity metrics, and gene annotations to prioritize rare, functionally significant variants. Additionally, TADA’s use of Bayesian false discovery rate (FDR) enhances power by balancing sensitivity and specificity in small-sample analyses, making it particularly suited to rare disorders like postpartum psychosis.

To elucidate the genetic basis of postpartum psychosis, this study investigates how common and rare genetic variation contributes to its risk. First, we estimate heritability using familial data from Swedish national registers. Next, we partition heritability by variant type using whole genome sequencing (WGS) data from the All of Us Research Program. Finally, we apply TADA to identify ultra-rare deleterious variants associated with postpartum psychosis and examine their overlap with genetic risk factors for schizophrenia, bipolar disorder, and multiple autoimmune disorders. Together, these analyses aim to clarify the genetic architecture of postpartum psychosis and provide a foundation for understanding its mechanisms and therapeutic targets.

## Results

Postpartum psychosis lacks diagnostic criteria within major classification systems, including the most recent version of the Diagnostic and Statistical Manual of Mental Disorders (DSM-5).^12^ Therefore, we employed the postpartum psychosis diagnostic classification used in our prior study and in existing literature.^10,13^ Specifically, we included mothers who received a diagnosis of mania, mixed psychotic depression, or psychotic episode within three months of their first live birth, a time frame that captures the typical onset window of the disorder.^1,14–16^ This definition was consistently applied to identify postpartum psychosis cases from both the Swedish national registers and the All of Us Research Program.

### Estimating postpartum psychosis heritability through familial concordance

To quantify the genetic contribution to postpartum psychosis, we first estimated heritability using familial concordance data from the Swedish national registers. This approach allowed us to assess overall narrow-sense heritability without relying on direct genetic data (Supplementary Table 1).

The study cohort included 1,648,759 mothers who experienced their first-ever childbirth between January 1, 1980, and October 31, 2017. Among these, 2,514 mothers (0.15%) were diagnosed with postpartum psychosis. Initially, we extracted pairs of siblings and cousins with a history of childbirth and calculated familial risk for postpartum psychosis in each relationship type (Methods). Then, to estimate heritability, we regressed the standard normal deviation of familial risk for postpartum psychosis against the standard normal deviation of population prevalence across degrees of relatedness. This analysis quantified the proportion of phenotypic variation attributable to additive genetic factors, effectively measuring narrow-sense heritability.

Our analysis indicated a heritability of 55% for postpartum psychosis (95% CI, 42-68%). This estimate aligns with heritability estimates for bipolar disorder, which range from 44% to 90%, depending on the study design.^11,17–19^

### Estimating postpartum psychosis heritability through common and rare genetic variation

Building on the family-based heritability estimate, we next quantified the contribution of both common and rare genetic variation to postpartum psychosis risk using WGS data. We analyzed WGS data from 301 mothers with postpartum psychosis and 1,505 ancestry-matched controls from the All of Us Research Program, a cohort representing diverse ancestral populations (Extended Data Fig. 1). We included variants with an allele frequency greater than 0.1% to assess the combined contribution of common and rare genetic variation to disease risk.

We estimated heritability using PCGC (Phenotype-Correlation Genotype-Correlation) regression, which operates on the liability scale and is well-suited for binary traits.^20^ This model is particularly effective for diverse populations because it does not rely on assumptions of uniform allele frequencies or linkage disequilibrium (LD) patterns across populations. Instead, PCGC explicitly models phenotype-genotype correlations, leverages pairwise relationships, and incorporates covariates such as principal components to account for population structure and stratification. Consistent with other studies, we adjusted for population structure using the first 50 principal components.^21–25^

Our analysis revealed that autosomal variants explain 36.9% ± 15.7% of the phenotypic variation for postpartum psychosis, providing an estimate of narrow-sense heritability based on inherited rare and common variation. The difference from the estimate derived from Swedish National records suggests that additional heritability may be attributable to ultra-rare variation, which was not captured in this analysis. Of interest, variants only on the X-chromosome explained 14.2% ± 8.3% of heritability, compared to 9.1% ± 8.3% for variants on chromosome 7, despite their comparable size,^26^ suggesting a role for X-linked genetic factors in the disorder.

### Comparing the burden of ultra-rare coding variation between cases and controls

While heritability estimates highlight the overall genetic contribution to postpartum psychosis, they do not identify specific genetic risk factors. To address this, we next investigated the burden of ultra-rare coding variants, which may play a critical role in the disorder’s genetic architecture due to their high potential functional impact.

We analyzed WGS data from 301 cases and 3,010 matched controls sourced from the All of Us Research Program. We defined ultra-rare variants as those present in < 0.1% of the population, with an allele count ≤ 5 in both our case-control cohort and the non-psychiatric subset of the gnomAD database (v3.1).^27^

We then compared the counts of autosomal protein-truncating variants (PTVs; including frameshift, stop gained, splice donor, splice acceptor, and transcript ablation variants), missense variants, and synonymous variants (SYNs) in protein-coding genes between cases and controls. Missense variants were further categorized by predicted impact using MPC (Missense Badness, PolyPhen-2, and Constraint) scores: MisB (MPC ≥ 2, highest probability of deleteriousness), MisA (1 ≤ MPC < 2), and Mis0 (MPC < 1, lowest probability of deleteriousness).^28^

The average total counts of each variant type were not significantly different between cases and controls in autosomes: PTVs (3.64 in cases vs. 3.50 in controls), MisB (1.45 vs. 1.41), MisA (9.62 vs. 9.83), Mis0 (59.61 vs. 59.80), and SYNs (39.32 vs. 39.24) (*P* > 0.05, binomial tests).

Given the lack of differences in overall counts, we examined whether variants were enriched in genes under stronger functional constraints as is observed in other psychiatric disorders.^29–33^ Using LOEUF (loss-of-function observed/expected upper bound fraction) score deciles, we grouped genes by constraint.^27^ To ensure sufficient statistical power for analysis, we combined deciles 1–3 as genes within deciles 1 and 2 contained too few variants for meaningful testing. We then assessed the burden of PTVs due to their high confidence of being loss-of-function. This approach revealed a significant enrichment of PTVs in cases within the most constrained genes (deciles 1–3) (*P* = 0.037, binomial test; Fig. 1).

**Fig. 1:**
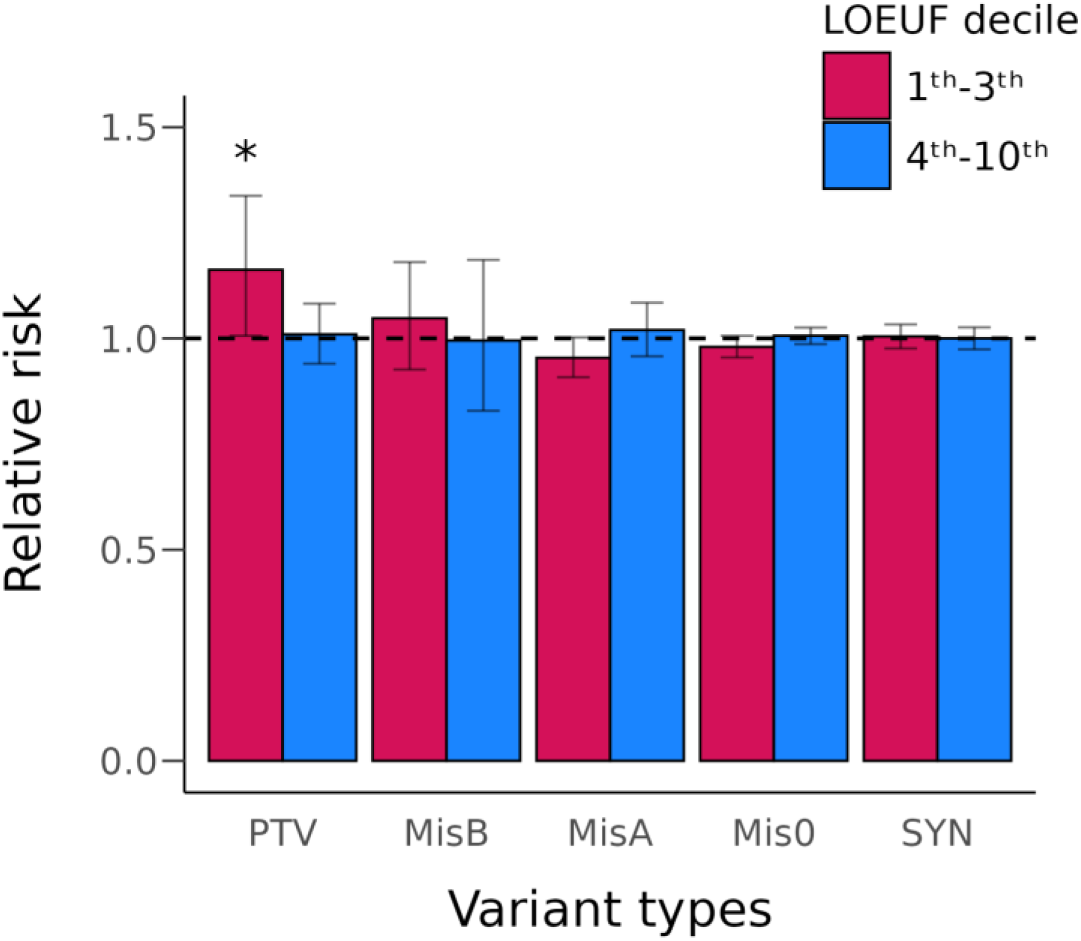
Distribution of ultra-rare autosomal protein-coding variants in cases and controls. The frequency of PTVs (Proteintruncating variants), missense variants and synonymous variants (SYNs) between cases and the unaffected control group in genes stratified by LOEUF (Loss-of-function observed/expected upper bound fraction) score. Missense variants were categorized using MPC (Missense badness, PolyPhen-2, and Constraint) scores (MisB: variants with MPC ≥ 2, MisA: variants with 2 > MPC ≥ 1, and Mis0: variants with 1 > MPC ≥ 0). \**P*<0.05 in binomial test.

To further validate the observed enrichment of PTVs in highly constrained genes (LOEUF deciles 1–3) and account for potential confounding factors such as population stratification, we performed a burden test using the sequential kernel association test for ordinal outcomes (SKAT-O).^34^ We analyzed the burden of ultra-rare PTVs in genes within the first to third LOEUF deciles, comparing cases and controls while adjusting for population subtle substructures with the first 50 principal components as covariates. This analysis confirmed that postpartum psychosis cases harbored a significantly elevated burden of PTVs in highly conserved genes (*P* = 0.005).

### Calculating the number of risk genes for postpartum psychosis

The observed enrichment of PTVs in conserved genes among postpartum psychosis cases underscores the potential to identify specific risk genes. As a first step toward systematically identifying these risk genes, we estimated the expected number of protein-coding autosomal genes contributing to postpartum psychosis risk that can be detected using sequencing data. To achieve this, we applied an approach inspired by the “unseen species problem,” commonly used in ecological studies,^35^ and in psychiatric genetics, to estimate risk gene counts.^36^ This approach infers the expected number of risk-associated genes, including those not yet observed, by analyzing the frequencies of deleterious *de novo* variants and estimating the likelihood that these variants represent a subset of the complete set of risk genes.

Because our cases lack parental data, we could not definitively distinguish inherited variants from *de novo* variants. To address this limitation, we examined the distribution of deleterious *de novo* PTVs and missense variants predicted to be damaging (MisB variants) in a cohort of 8,378 trio offspring.^37^ This distribution was then used to randomly assign de novo status to variants in our case-control dataset across 1,000 permutations. For each permutation, we estimated the expected number of risk genes that could be identified through sequencing (see Extended Data Fig. 2 and Methods).

The simulations yielded a mean estimate of 84 risk genes, with a 95% confidence interval ranging from 68 to 102. This represents approximately 0.46% of the 18,128 autosomal protein-coding genes assessed. These estimates should be refined as additional sequencing data become available. Nonetheless, this estimate of 84 risk genes is substantially lower than corresponding estimates for other psychiatric conditions (e.g., autism, which is estimated to involve 1,000 risk genes).^36^ The smaller pool of risk genes implicated in postpartum psychosis mitigates the smaller sample size by increasing the likelihood of observing deleterious variants in the same risk gene among cases.

### Identification of postpartum psychosis risk genes

We next sought to identify specific risk genes for postpartum psychosis. We implemented the TADA test, a widely adopted approach for complex disorders that integrates rare loss-of-function variations across multiple variant classes, taking into account evolutionary constraints and the above estimated number of risk genes.^36^ For each autosomal gene, we calculated the strength of association, considering different types of variants and prior probabilities of relative risk (Supplementary Table 2). Across all tested genes, we did not observe inflated significance values, as evidenced by a genomic inflation factor (λ_GC_) of 1.002 and a standard quantile-quantile distribution of *P* values (Extended Data Fig. 3). Our analysis identified *DNMT1* (DNA Methyltransferase 1) and *HMGCR* (3-Hydroxy-3-Methylglutaryl-CoA Reductase) as high-confidence risk genes with q value < 0.1. This threshold is widely used across psychiatric genetics.^32,33,38,39^ In addition, we found three potential risk genes with 0.1 ≤ q value < 0.3 (Table 1 and Fig. 2). Under the All of Us Data and Statistics Dissemination Policy, we are unable to display the ultra-rare variant count per gene used in the TADA test.

**Fig. 2:**
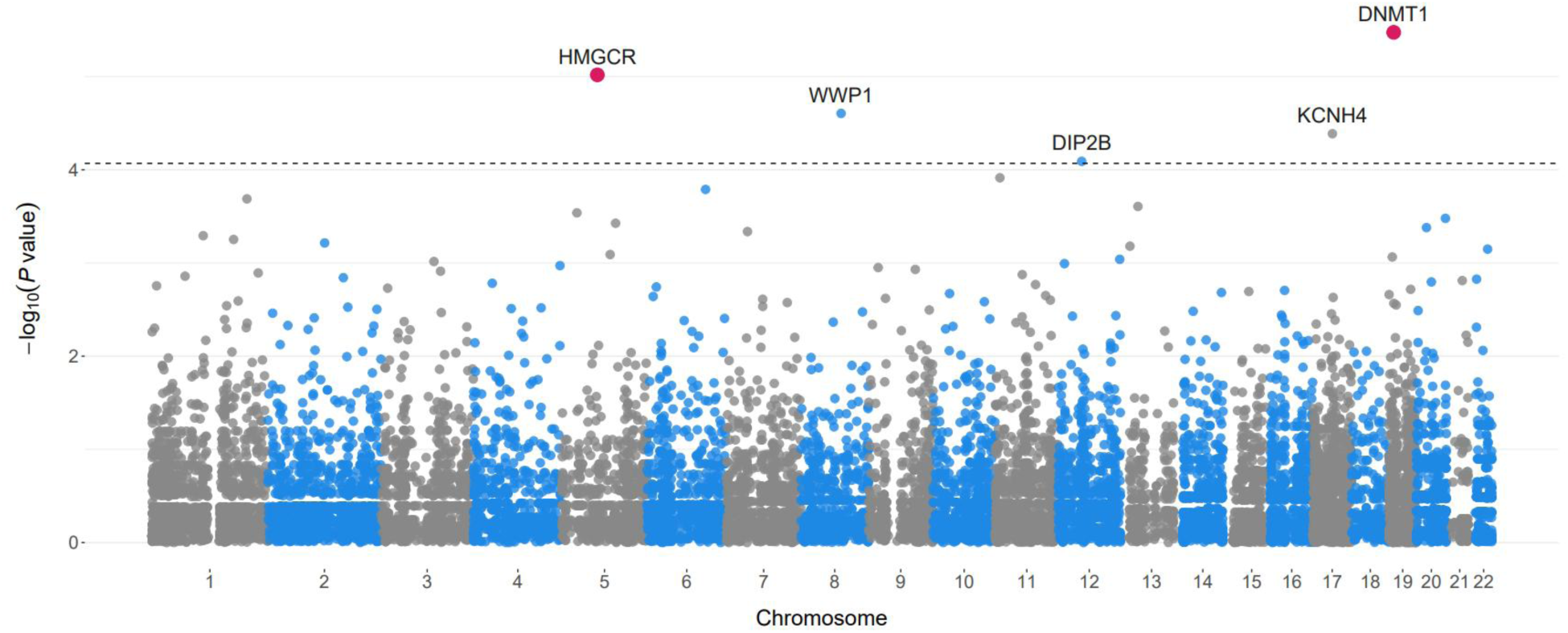
Association of protein-coding genes to postpartum psychosis risk. Points represent the *P* value association of individual protein-coding genes in the postpartum psychosis TADA result. Genes with a q value < 0.1 (red points) or < 0.3 (dotted line) are labeled.

**Table 1.** Genes with q value <0.3 in TADA result.

| Gene | pLI | LOEUF | q value* | P value<sup>#</sup> |
| --- | --- | --- | --- | --- |
| ***DNMT1*** | **1.000** | **0.143** | **0.060** | **$3.36 \times 10^{-6}$** |
| ***HMGCR*** | **0.999** | **0.260** | **0.086** | **$9.59 \times 10^{-6}$** |
| *WWP1* | 1.000 | 0.248 | 0.149 | $2.49 \times 10^{-5}$ |
| *KCNH4* | 1.000 | 0.215 | 0.185 | $4.10 \times 10^{-5}$ |
| *DIP2B* | 0.999 | 0.266 | 0.293 | $8.12 \times 10^{-5}$ |

LOEUF, loss-of-function observed/expected upper bound fraction score; pLI, the probability of being loss-of-function intolerant score; TADA, transmission and *de novo* association.
\* q value: false discovery rate, calculated in transmission and *de novo* association test.
<sup>#</sup>P value: P value of transmission and *de novo* association test.

### Association between ultra-rare variants in *DNMT1* and *HMGCR* and psychiatric disorders

We sought to determine whether ultra-rare deleterious variants (PTV and MisB) in *DNMT1* and *HMGCR* were also associated with a broader spectrum of psychiatric disorders. This analysis aimed to assess the broader relevance of these genes in shared neuropsychiatric pathways and to indirectly validate their biological significance across psychiatric conditions beyond postpartum psychosis.

To conduct this analysis, we utilized 240,009 samples from the All of Us database, excluding cases of postpartum psychosis, to ensure the findings were independent of the primary study cohort. Psychiatric cases were identified using ICD-10 diagnostic codes, while controls were defined as individuals lacking such codes. We applied the SKAT-O method to assess the burden of ultra-rare deleterious variants, incorporating sex and the first ten principal components to account for broad ancestry and population structure. A Bonferroni correction was used to adjust for multiple testing across ICD-10 codes.

Our analysis identified significant associations between *DNMT1* and 12 psychiatric disorders, with the strongest signals observed for emotional disorders with onset specific to childhood, mental and behavioral disorders due to psychoactive substance use involving hallucinogens, and mental and behavioral disorders associated with the puerperium (Supplementary Table 3). For *HMGCR,* we identified significant associations for five psychiatric disorders, including mental disorder not otherwise specified, mental and behavioral disorders due to multiple drug use and other psychoactive substances, and dementia in other diseases classified elsewhere. Together, these results suggest *DNMT1* and *HMGCR* are relevant to psychiatric conditions and support their role in postpartum psychosis.

### Comparative analysis of postpartum psychosis and other disorders

Existing studies suggest a close link between postpartum psychosis and both bipolar disorder and schizophrenia.^40–48^ Furthermore, growing evidence indicates that immune system changes related to autoimmune disorders during and after pregnancy may contribute to the development of postpartum psychosis.^49^ We, therefore, investigated the proportion of risk genes for bipolar disorder, schizophrenia, and 29 autoimmune diseases (the latter chosen based on data availability, see Methods) identified from WGS studies that show evidence of association in the postpartum psychosis TADA result using the “propTrueNull” function in the *limma* package in R (Methods).^30,31,50^ Briefly, this approach compares per-gene *P* values and estimates the proportion of true null hypotheses in the joint, multiple-testing scenario. While individual genes might not achieve significance in isolation, this method identifies their collective impact, particularly when genes share related pathways or biological functions.

We observed that 17% of the top 200 risk genes for bipolar disorder (*P*=0.019), 21% for schizophrenia (*P*=0.023), 16% for rheumatoid arthritis (*P*=0.046), 25% for myasthenia gravis (*P*=0.010), 20% for Sjogren’s syndrome (*P*=0.022), and 18% for Crohn’s disease (*P*=0.022) exhibit a possible association with postpartum psychosis (Fig. 3a-f and Supplementary Table 4). Notably, *HMGCR* (postpartum psychosis *P*=9.59×10^-6^) emerged as one of the top 200 risk genes for both schizophrenia (*P*=6.44×10^-4^) and rheumatoid arthritis (*P*=3.87×10^-3^) (Fig. 3a and b). Similarly, *WWP1* (WW domain containing E3 ubiquitin protein ligase 1) (postpartum psychosis *P*= 2.49×10^-5^) was also a bipolar disorder risk gene (*P*=6.52×10^-3^) (Fig. 3c). These findings suggest that some risk genes for postpartum psychosis share genetic etiology with other psychiatric disorders, while others, such as *DNMT1*, may represent distinct genetic risk factors specific to postpartum psychosis.

**Fig. 3:**
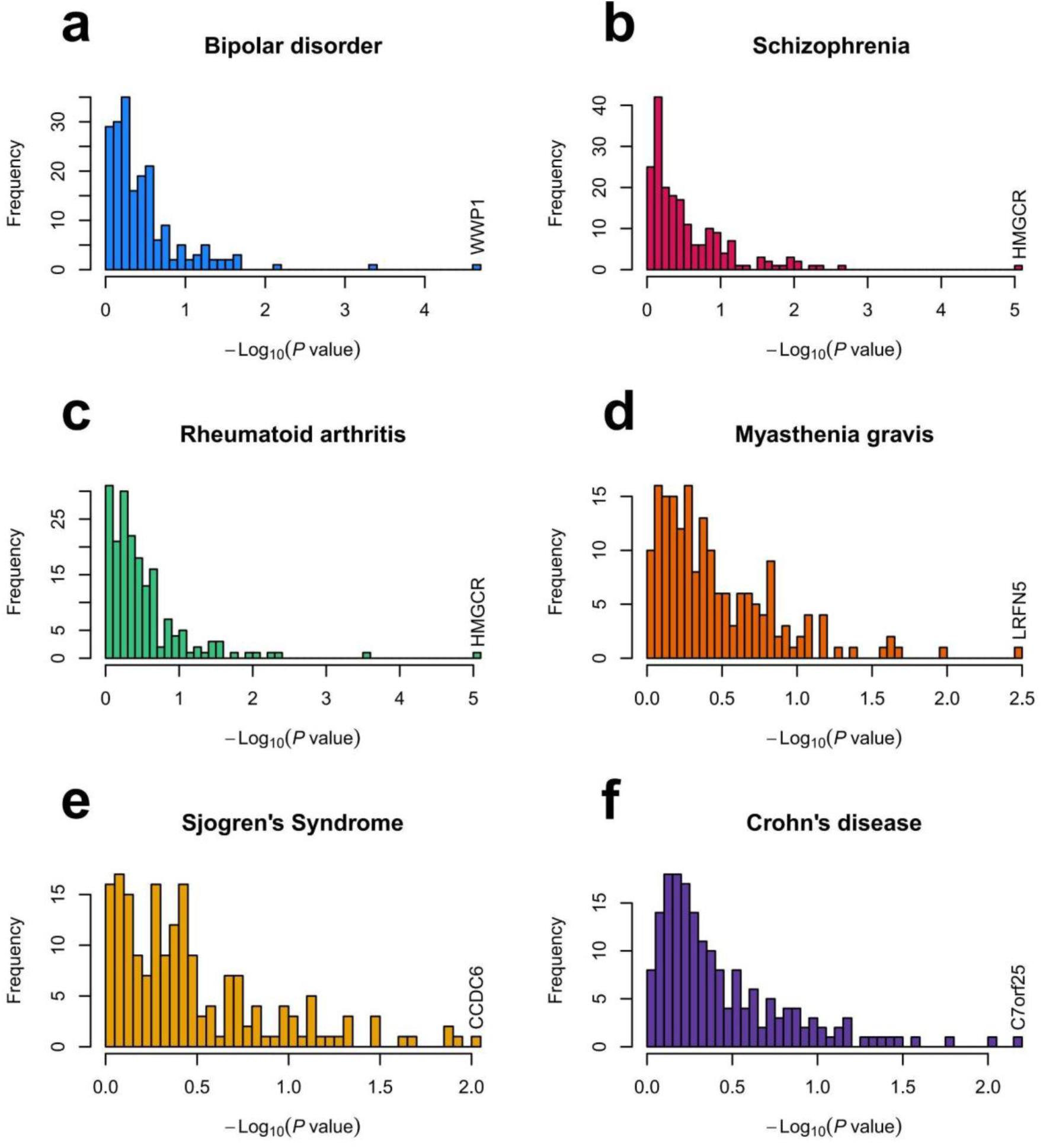
Association of postpartum psychosis risk genes in other psychiatric and autoimmune disorders. **a)** The association (TADA *P* value) distribution of the top 200 bipolar disorder-associated genes from Palmer et al. 2022 for postpartum psychosis. **b)** As in panel ‘a’ for the top 200 schizophrenia-associated genes from Singh et al. 2022 **c)** As in panel ‘a’ for the top 200 rheumatoid arthritis-associated genes from Zhang et al. 2024. **d)** As in panel ‘a’ for the top 200 myasthenia gravis-associated genes from Yang et al. 2024. **e)** As in panel ‘a’ for the top 200 Sjogren’s syndrome-associated genes from Yang et al. 2024. **f)** As in panel ‘a’ for the top 200 Crohn’s disease-associated genes from Yang et al. 2024. In each panel, a gene with the most significant *P* value was labeled.

## Discussion

The genetic etiology of postpartum psychosis has hardly been investigated, even though it occurs in 1 out of 1000 new mothers and 1 out of 5 mothers with bipolar disorder. To deepen our understanding, we employed two complementary approaches to estimate the heritability of this disorder. First, a family-based analysis using data from the Swedish national registers estimated the heritability of postpartum psychosis to be 55% (95% CI: 42%–68%). This estimate primarily reflects autosomal genetic contributions, as the relatedness matrix used in such models does not typically include sex chromosome inheritance. However, the inclusion of female sibling and cousin relationships suggests some contribution from X-linked factors. Unique features of X chromosome inheritance, such as X-inactivation in females and the absence of recombination in males, complicate the incorporation of X-linked contributions into standard heritability models, which are primarily designed to estimate autosomal effects.

We additionally assessed heritability using common and rare genetic variation using WGS data from All of Us participants, revealing a comparable but lower autosomal heritability estimate of 37% (SE: 16%). Notably, this analysis identified a contribution from the X chromosome, highlighting its potential role in the disorder’s genetic risk. The disparity between heritability estimates derived from family-based and genomic data underscores the concept of “missing heritability”. Family-based methods estimate total heritability, capturing contributions from rare and family-specific variants, non-coding regions, and partially account for complex genetic effects such as epistasis and X-linked inheritance. In contrast, WGS-based approaches may underestimate heritability due to limited power to detect all relevant variation. Environmental contributions shared among relatives may also inflate family-based estimates.

We next examined the contribution of ultra-rare coding variants to the genetic risk of postpartum psychosis. Our analysis revealed that ultra-rare coding variants, particularly PTVs, significantly influence the risk of postpartum psychosis. The burden of ultra-rare PTVs was significantly elevated in highly conserved genes among postpartum psychosis cases. Similar enrichment of ultra-rare PTVs in conserved genes has been observed in other psychiatric disorders, including bipolar disorder, schizophrenia, autism spectrum disorder, attention-deficit/hyperactivity disorder, obsessive-compulsive disorder, Tourette disorder, and major depressive disorder, also show significant enrichment of ultra-rare PTVs in conserved genes.^29–31,33,38,51–54^

Building on the contribution of ultra-rare coding variants to the genetic risk of postpartum psychosis, we estimated that 68 to 102 autosomal risk genes may contribute to the disorder, identifiable through sequencing data. The role of X chromosome risk genes remains an avenue for future exploration as crucial metrics of constraint are not available for these genes.

Among autosomal genes, we identified *DNMT1* and *HMGCR* as high-confidence risk genes with a false discovery rate (FDR) < 0.1, a stringent threshold commonly used in similar rare variant studies to reduce false discoveries.^29,32,33,38,39^ *DNMT1* is known for its role in preserving genomic methylation patterns through maintenance methylation rather than initiating new methylation marks.^55,56^ Alterations in *DNMT1* function have broad pleiotropic effects, and full loss of function is generally nonviable.^57^ However, neuron-specific conditional expression studies suggest roles for *DNMT1* in synaptic plasticity and neuronal differentiation.^57–59^ Observational studies in humans have reported elevated *DNMT1* expression in patients with bipolar disorder and schizophrenia, although the mechanistic importance of this observation remains unclear.^60,61^ *DNMT1* deficiency also disrupts immune homeostasis by altering DNA methylation and the function of critical immune cells, such as plasmacytoid dendritic cells (pDCs).^62^

*HMGCR* encodes the rate-limiting enzyme in cholesterol biosynthesis.^63,64^ Low serum cholesterol is known to be associated with both first-onset psychosis^65^ and suicidal behavior.^66,67^ Potential mechanisms include alterations in neuronal membrane viscosity, affecting synaptic transmission, or neuroinflammatory effects resulting from altered composition of lipid rafts.^67^ In addition, a recent schizophrenia genome-wide association study identified a variant (rs113537149, *P*=2.44×10^-7^), located 52 kb upstream of *HMGCR*.^68^ and within the linkage disequilibrium block encompassing *HMGCR*.

The role of *HMGCR* in postpartum psychosis may also be understood in the context of pregnancy-associated metabolic changes. Pregnancy induces substantial alterations in lipid metabolism, including marked increases in serum cholesterol levels over the course of pregnancy, which rapidly return to baseline level postpartum. Cholesterol serves as a backbone for steroid hormone synthesis, and the extreme hormonal fluctuations associated with childbirth and the postpartum period may interact with disruptions in cholesterol biosynthesis, potentially contributing to the pathophysiology of postpartum psychosis.

Beyond identifying specific risk genes for postpartum psychosis, our findings highlight shared genetic architecture with other psychiatric and autoimmune disorders. In particular, the overlap with bipolar disorder and schizophrenia risk genes emphasizes the clinical and genetic parallels with postpartum psychosis, as well as with autoimmune conditions that carry increased risk during the postpartum period. Notably, *HMGCR* emerges as a significant risk gene for postpartum psychosis, schizophrenia, and rheumatoid arthritis, suggesting potential dysregulation in metabolic or immune pathways contributing to the pathogenesis of these conditions. The postpartum period is characterized by immune reactivation and hormonal fluctuations, which may exacerbate risks for autoimmune diseases. For example, the risk of Myasthenia gravis onset risk is five times higher in the first six months postpartum compared to during pregnancy,^69^ and women with Crohn’s disease are more likely to experience disease flare-ups and new-onset psychiatric diagnoses postpartum.^70,71^ Similarly, rheumatoid arthritis has been linked to an increased risk of postpartum psychiatric disorders in women,^72^ reinforcing the interconnected roles of immune dysregulation and mental health during this critical period.

Although postpartum psychosis cases were limited, this study validated its findings using multiple complementary approaches. Strengths include (1) The use of a population-based sample from the Swedish national registers minimizes self-selection bias, ensuring broader representation compared to studies relying on small clinical cohorts, which are often affected by pre-existing psychiatric diagnoses. (2) The combination of family-based heritability estimates with WGS data from the All of Us Research Program provides a complementary approach to understanding the genetic architecture of postpartum psychosis. (3) The inclusion of X chromosome contributions in the heritability analysis, often overlooked in genetic studies, is particularly relevant for a condition exclusive to females. (4) The study highlights ultra-rare coding variants, such as protein-truncating variants, and identifies specific risk genes like *DNMT1* and *HMGCR*, offering potential targets for functional research and therapeutic development. (5) The exploration of genetic overlaps with other psychiatric and autoimmune disorders provides broader insights into shared pathways that may contribute to postpartum psychosis and related conditions. Together, these strengths position this study as a significant contribution to the under-researched field of postpartum psychosis genetics.

While this study provides valuable insights into the genetic risk factors for postpartum psychosis, it has several limitations. (1) Postpartum psychosis is a broad term, and its definition may vary across studies and databases. This variability could affect the comparability of our findings with other research. (2) The family-based analysis focused on families with female pairs, potentially underrepresenting the full spectrum of familial risk, particularly in families with only one female sibling. (3) The number of control females with a history of childbirth was insufficient for precise case-control matching. As a result, the control group included some females with unknown postpartum psychosis status. Although the low incidence of postpartum psychosis makes this statistically unlikely, the inclusion of undiagnosed cases as controls could bias the results toward the null hypothesis, potentially reducing the ability to detect significant associations. (4) We were underpowered to assess genetic risk across different ancestries. Future research should prioritize ancestry-specific analyses to uncover potential genetic variations in risk among diverse populations, enhancing the generalizability of our findings. (5) Given the novelty of our findings and the limited data available, replication is necessary to confirm the accuracy of our genetic associations, address potential false positives and negatives, and mitigate overfitting. The low prevalence of postpartum psychosis, combined with the lack of comprehensive diagnostic information, limited our ability to source additional samples from the UK Biobank or other databases. (6) A limitation of the comparative analysis is the selection of the top 200 genes from existing studies, even though not all are statistically significant. This approach may introduce biases, as it includes genes with weaker evidence, diluting the overall signal and reducing power to detect true overlaps. Additionally, predefined gene sets may lead to false positives, exclude relevant genes, or reflect limitations of prior studies, such as small sample sizes or unrepresentative populations. These constraints may limit the generalizability of our findings if genetic architectures differ across populations. (7) We did not account for participants’ psychiatric history, such as distinguishing postpartum psychosis as a continuation of a pre-existing disorder from new-onset cases following childbirth. Given that many individuals with postpartum psychosis have a history of other psychotic disorders, our findings may also reflect general patterns of severe mental illness rather than postpartum psychosis-specific factors only. Future research should stratify participants by psychiatric history to improve specificity and better understand overlaps with other psychiatric conditions. (8) Under the All of Us Data and Statistics Dissemination Policy, we were unable to report ultra-rare variant counts per gene, limiting the transparency of our results.

Finally, for heritability estimation, we included the first 50 PCs to account for subtle population structure. Rare variant association analysis using TADA, however, presents additional complexities, as TADA does not support direct ancestry adjustment. One potential solution would be to stratify the population into major ancestry groups, estimate TADA parameters separately for each group, and perform TADA analyses within each group to compute Bayes factors. These Bayes factors could then be combined multiplicatively to obtain overall Bayes factors. However, this stratification approach has limitations, particularly for admixed populations with highly heterogeneous genetic structures, as it does not fully capture the complexity of local ancestry. In our analysis, we addressed ancestry by matching cases and controls based on the first five PCs, effectively accounting for major axes of genetic variation. This matching approach indirectly adjusts for ancestry and minimizes confounding due to population stratification. While this method reduces bias, future works could incorporate local ancestry inference to further refine parameters for admixed populations.

In summary, this study offers important insights into the genetic architecture of postpartum psychosis, emphasizing the significant contributions of both common and rare genetic variants to its risk. Further studies using larger will be essential to validate these findings and identify additional genetic factors. Functional analyses of identified risk genes, such as *DNMT1* and *HMGCR*, will be critical to uncover their roles in the disorder and explore therapeutic potential. Furthermore, investigating interactions between genetic variants and environmental factors, such as hormonal fluctuations during and after pregnancy, could provide deeper insights into the mechanisms underlying postpartum psychosis.

## Methods

### Study population: Swedish national registers

We utilized a cohort from the Swedish national registers to estimate the heritability of postpartum psychosis, consistent with our prior analysis of familial recurrence for this disorder. The study included data from the Medical Birth Register with a first-ever live birth between January 1, 1980, and October 31, 2017, with follow-up available until December 31, 2017. Female full siblings and first-degree cousins were identified using the Multi-Generation Register. Psychiatric disorder diagnoses were obtained from the Swedish National Patient Register, which includes both inpatient and outpatient care data. Ethical approval and a waiver of informed consent were granted by the Regional Ethical Review Board in Stockholm, Sweden.

To identify cases of postpartum psychosis, we used ICD-8, ICD-9, and ICD-10 codes to capture all instances of mania and/or psychosis with onset within 0–3 months postpartum following the first live birth. Diagnoses included “brief psychotic disorder,” “psychotic disorders not due to a substance or known physiological conditions,” “manic episode,” “bipolar disorder,” “major depressive disorder, single episode, severe with psychotic features,” “recurrent depressive disorder, current episode severe with psychotic features,” and “puerperal psychosis.” A detailed list of the specific ICD codes is provided in Supplementary Table 5.

### Calculating heritability from familial concordance

We used a family-based quasi-experimental design that leverages inherent differences in family relationships, such as variations in genetic relatedness and environmental exposure within the same household. This approach allows us to observe the effects of genetic and environmental factors in a quasi-controlled setting, closely mimicking the conditions of randomized controlled trials. Using the pairs of siblings and cousins, we applied Falconer’s Liability Threshold Model to quantify the direct additive genetic effect in postpartum psychosis. Initially, we computed the familial risk (FR_Postpartum psychosis) — the likelihood that a proband with postpartum psychosis has an affected relative — across the different relationship types. Using formula 25.1.a from Chapter 25 (pages 730-736) of (1), we estimated the regression coefficient (b) between familial risk in relatives of probands with postpartum psychosis and population prevalence for each relationship category:

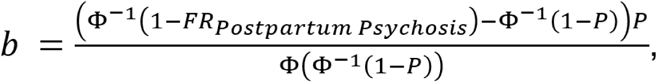

where P represents the population prevalence of postpartum psychosis, the prevalence of the trait among various types of relationships often mirrors the population prevalence. If this alignment does not occur, we adjust by using the prevalence rate specific to each relationship type to calculate the regression coefficient for that type. Φ and Φ^(−1) denote the standard normal distribution and its inverse, respectively. The regression coefficient (b) for each relationship type is equated to its expected contribution from direct additive genetics to the postpartum psychosis phenotype (Supplementary Table 6). Subsequently, weighted least squares are employed to estimate the variance components, with weights proportional to the number of probands in each relationship type (Supplementary Table 7).

### Study population: All of Us

Within the All of Us Research Program (controlled tier, v7), we identified a total of 431 samples with postpartum psychosis. Of these, 268 female samples were classified under the ICD-10 code F53.1 (postpartum psychosis). An additional 163 cases were identified based on the definition used for postpartum psychosis in the Swedish registers. This definition included diagnoses of brief psychotic disorder, psychotic disorders not due to a substance or known physiological condition, manic episodes, bipolar disorder, severe major depressive disorder with psychotic features (single episode or recurrent), and puerperal psychosis, occurring within 0–3 months of childbirth (Supplementary Table 8).

We defined 170,161 female samples without any SNOMED codes for mental disorders as controls. Case-control matching was performed using the R package ‘optmatch’, which facilitates optimal matching in observational studies to minimize bias and improve causal inference. Distances between units were calculated using the Euclidean distance metric, and matching was based on principal components (PCs) 1–5, derived from WGS data provided by the All of Us Research Program. From this process, 10,000 candidate ancestry-matched controls were selected. We used 1–5 PCs for the matching as these PCs capture genetic variation due to broad ancestry differences, ensuring effective matching without over-adjusting.

### Quality control of whole genome sequencing data

We analyzed short-read whole genome sequencing data from 310 cases and 9,001 controls from All of Us. Alignment uses the GRCh38dh reference genome. Following All of Us guidelines, ^73^ we excluded 31 individuals with <2.4M or >5.0M total variants, >100k variants not in gnomAD v3.1,^27^ or a heterozygous-homozygous variant ratio >3.3. Additionally, we removed 1,039,352 low-quality variants (GQ ≤ 20, DP ≤ 10, and AB ≤ 0.2 for heterozygotes, ExcessHet <54.69, QUAL <60 for SNVs and <69 for short InDels). Our quality control excluded 43 samples with call rates >3 standard deviations below the mean, 189 individuals with close genetic relatedness of kinship value >0.1 in the Kinship-based INference for Genome-wide association studies (KING),^74^ and 7 sex-mismatched individuals. After filtering, 305 cases and 8,736 controls were retained. We further filtered out homozygous reference calls with <90% read depth supporting the reference allele or GQ < 25; heterozygous calls with an allele balance <30%, GQ < 25, or >1×10⁻⁹ probability of the allele balance based on a binomial distribution centered on 0.5; and homozygous alternate calls with <90% read depth supporting the alternate allele or GQ < 25. Variants with a call rate ≤90% and Hardy-Weinberg equilibrium *P* < 1×10⁻¹² were also removed.

We used the Variant Effect Predictor (VEP) to predict the potential functional effect of variants in exons.^75^ We classified variants into protein-truncating variants (PTVs: frameshift, stop gained, splice donor, and splice acceptor), missense, and synonymous variants. For PTVs, we applied LOFTEE (Loss-Of-Function Transcript Effect Estimator), requiring an ‘HC’ designation and no LOFTEE flags except ‘SINGLE_EXON.’ Missense variants were categorized by MPC scores: MisB (MPC ≥ 2), MisA (2 > MPC ≥ 1), and Mis0 (1 > MPC ≥ 0).^28^

For annotating ultra-rare variants, we included variants with an allele count of ≤ 5 in both the case-control cohort and the non-psychiatric subset of the gnomAD database (v3.1).^27^ We examined the distribution of rare synonymous variants, removing 4 cases and 108 controls with counts >3 standard deviations above the mean. After quality control, there were 301 cases and 8,628 controls.

### Calculating heritability with whole genome sequencing data

We estimated heritability using PCGC (Phenotype-Correlation Genotype-Correlation) regression, an alternative to restricted maximum likelihood (REML) for heritability estimation in binary traits, such as diseases.^20^ PCGC estimates heritability on the liability scale, which is more suitable for binary traits, and addresses ascertainment bias by accounting for the bimodal liability distribution and phenotype-covariate correlations introduced by case over-sampling. To calculate heritability using both common and rare variants from whole genome sequencing data, we retained variants with an allele frequency greater than 0.1% in the matched case-control dataset. We conducted 1:5 case-control matching using ‘optmatch’ based on the first five PCs that we calculated using the quality controlled whole genome sequencing data, yielding 301 cases and 1,505 matched controls. We applied a prevalence of 0.15% and included the first 50 PCs as covariates to comprehensively adjust for population structure, particularly when dealing with whole genome sequencing data, where subtle population substructure can influence heritability estimates. In this study, we treated the X chromosome as equivalent to autosomes for several reasons. First, our analysis included only female samples, thereby eliminating the differences in copy number between sexes that typically necessitate adjustments for the X chromosome. In females, both X chromosomes are present, and the genetic variance attributable to X-linked loci can be modeled similarly to autosomal loci. Second, by restricting our dataset to a single sex, we bypassed the need to account for dosage compensation mechanisms, as these primarily address differences between males and females.

### Calculating the number of risk genes

As our data are limited to case-control studies, we first examined how the number of ultra-rare deleterious *de novo* variants (PTVs and MisB variants) are distributed among 8,378 unaffected siblings from the SPARK dataset.^37^ For the ultra-rare variant study, we performed 1:10 case-control matching using ‘optmatch,’ as described above, based on the first five PCs that we calculated using the quality controlled whole genome sequencing data. This increased the control sample size, resulting in 301 cases and 3,010 matched controls. We then randomly selected ultra-rare deleterious variants from our case-control dataset to be *de novo*, ensuring they matched the distribution observed in the SPARK data. We proceeded under the assumption that the number of deleterious variants in cases minus those in controls (d) contributes to the risk of postpartum psychosis, and genes exhibiting recurrent variants (x) were regarded as indicative of risk-associated events.

We performed 1,000 *de novo* sampling permutations and applied the formula to calculate the number of risk genes in each permutation (C): C = c/u + g^2 * d * (1-u)/u, in which: c as the total number of distinct genes with a *de novo* (d - x), c1 as the count of singleton genes (d - 2x), g as the coefficient of variation in the fractions of genes of each type, and u as 1 – c1/d.^76^ This calculation was utilized to estimate the number of genes contributing autism spectrum disorder risk.^35^ For our calculations, we assume g to be one due to the limited number of observations. We calculated a mean and 95% confidence intervals based on the result of 1,000 permutations.

### Identification of risk genes

We used the Transmission and De Novo Association Test (TADA) to identify risk genes for postpartum psychosis.^36^ We employed the extended TADA method to integrate rare coding variants (PTVs, MisB, MisA, Mis0, and synonymous variants) and LOEUF scores.^27,29,38^ Our TADA variant count tallied the number of rare case-control variants for each gene and the LOEUF score for each gene (Supplementary Table 3). According to the All of Us Data and Statistics Dissemination Policy, we are unable to display the ultra-rare variant count for each gene.

TADA calculates a Bayesian Factor (BF) to quantify the statistical strength of gene-level association. This calculation is based on various inputs, including the count of variant events, sample size, proportion of risk genes, and a prior assessment of variant risk (gamma) within each gene. For each gene, gamma for PTVs, gamma for MisB variants, and gamma for MisA variants were calculated as “the relative risk of the variant” divided by “the estimated fraction of genes that substantially influence postpartum psychosis risk,” smoothed over the LOEUF score. We created bins of data ordered by LOEUF and fitted a logistic curve to the data. We then computed a rolling-average gamma for PTVs, MisB variants, and MisA variants for the fraction of risk genes, representing the relative enrichment in cases compared to controls (Extended Data Fig. 4a and b, and Supplementary Table 3). As the relative risk of MisA variants was lower than 1, we set the gamma MisA to 1 (no risk).

To calculate the total gene-level BF, we multiplied the BFs of the three variant classes. To ensure that evidence for association from one variant type is not diluted by evidence against association from another variant type, we set a minimum BF of 1 for each variant class. We then converted BFs to posterior probabilities, which were used to compute *P* values and q values for adjusting multiple testing, generating a list of candidate risk genes. We defined risk genes by applying widely used statistical thresholds: genes with an FDR below 0.3 were categorized as ‘potential’ risk genes, while those with an FDR below 0.1 were considered ‘high-confidence’ risk genes.^32,33,38,39^

### Ultra-rare variant burden in *HMGCR* and *DNMT1* and related psychiatric disorders

We investigated which psychiatric disorder is associated with the burden of ultra-rare deleterious variants in *HMGCR* or *DNMT1*. The study cohort consisted of 240,009 samples with WGS data from the All of Us research database. Notably, samples with postpartum psychosis were not included in this study cohort. For phenotype classification, cases were defined as samples with an ICD-10 code beginning with “F”, which is the mental disorder, while controls were those without the specific ICD-10 code. For instance, if there were 300 samples with the F01 code, the remaining 240,019 samples (240,319 total - 300 cases) served as controls. The genetic data used in this analysis included a genotype matrix of ultra-rare deleterious variants, specifically PTV or missense variants predicted to be damaging (MisB), in *HMGCR* or *DNMT1*. We conducted a burden test using SKAT-O, incorporating sex and the first ten PCs (PC1-10) as covariates to account for ancestry.^34^ A threshold for statistical significance was set at a p-value of 0.00034 (0.05/144), accounting for the 72 ICD-10 codes beginning with “F” and the two genes tested.

### Comparative analysis

To assess what proportion of risk genes for bipolar disorder, schizophrenia, and rheumatoid arthritis show evidence of association in postpartum psychosis, we selected the top 200 genes from existing whole-exome sequencing studies.^30,31,50^ We choose 29 autoimmune diseases (alopecia areata, ankylosing spondylitis, asthma, autoimmune hepatitis, autoimmune hypothyroidism, Behcet’s disease, celiac disease, Crohn’s diease, Graves’ disease, Guillain-Barre syndrome, idiopathic thrombocytopenic purpura, lichen planus, multiple sclerosis, myasthenia gravis, myositis, narcolepsy, necrotizing vasculopathies, pernicious anaemia, primary biliary cirrhosis, psoriasis, psoriatic and enteropathic arthropathies, rheumatoid arthritis, sarcoidosis, scleroderma, Sjogren’s syndrome, systemic lupus erythematosus, type I diabetes mellitus, ulcerative colitis, vitiligo) based on availability.^50,77^ The purpose of selecting the top 200 genes is to focus on candidates that have shown some level of association or biological relevance in other studies, even if not all of them reach strict statistical significance thresholds. Genes may not be individually statistically significant but could still contribute meaningfully when considered as a group, especially if they are involved in similar pathways or biological processes. We compared significance using the “propTrueNull” function in the R limma package. The proportion of true null hypotheses (π0) was calculated using a method of “convest” in the propTrueNull function. The proportion of alternative hypotheses (π1) was estimated based on 1 – π0, representing the proportion of risk genes. A π1 >0 indicates that there is a significant portion of hypotheses that are likely true alternatives, suggesting that there are overlapping risk genes between postpartum psychosis and other disorders. To assess the statistical significance of risk gene overlap between postpartum psychosis and the second psychiatric/autoimmune disorder, we performed 1000 iterations of random selection from the total list of autosomal protein-coding genes. In each iteration, we calculated π1 and derived an empirical *P* value for the observed overlap.

## Data Availability

Data may be obtained from a third party and are not publicly available. Data cannot be shared publicly owing to restrictions by law. Data are available from the National Medical Registries in Sweden after approval by the Swedish Ethical Review Authority.
Data from All of Us can be access through their website.

## Acknowledgments

This study was supported by a grant from the Beatrice and Samuel A. Seaver Foundation (BM, APK); the National Institute of Mental Health (NIMH), R21MH131933 (VB, KH, APK, BM, TMO, TKR), R01 HD111117 (TKR); Out to Innovate Career Development Fellowship (APK); and the 2020 NARSAD Young Investigator Grant from the Brain & Behavior Research Foundation (BM, grant number 29355). The All of Us Research Program is supported by the National Institutes of Health, Office of the Director: Regional Medical Centers: 1 OT2 OD026549; 1 OT2 OD026554; 1 OT2 OD026557; 1 OT2 OD026556; 1 OT2 OD026550; 1 OT2 OD 026552; 1 OT2 OD026553; 1 OT2 OD026548; 1 OT2 OD026551; 1 OT2 OD026555; IAA #: AOD 16037; Federally Qualified Health Centers: HHSN 263201600085U; Data and Research Center: 5 U2C OD023196; Biobank: 1 U24 OD023121; The Participant Center: U24 OD023176; Participant Technology Systems Center: 1 U24 OD023163; Communications and Engagement: 3 OT2 OD023205; 3 OT2 OD023206; and Community Partners: 1 OT2 OD025277; 3 OT2 OD025315; 1 OT2 OD025337; 1 OT2 OD025276. In addition, the All of Us Research Program would not be possible without the partnership of its participants.

## Author contributions

Study concept and design: VB, SJ, BM

Acquisition, analysis, or interpretation of data: VB, MC, KH, SJ, BM, TMO, TKR Drafting of the manuscript: VB, MC, SJ, BM, TKR

Critical revision of the manuscript for important intellectual content: All authors Statistical analysis: SJ, BM

Obtained funding: VB, BM Study supervision: VB, BM

## Code availability

Code and resources used in this study is available on GitHub at (https://github.com/MahjaniLab/pp_WGS). For any further inquiries or requests for code not available in the repository, please contact the corresponding author.

