## Extended data for "Genetic Architecture of Postpartum Psychosis: From Common to Rare Genetic Variation"

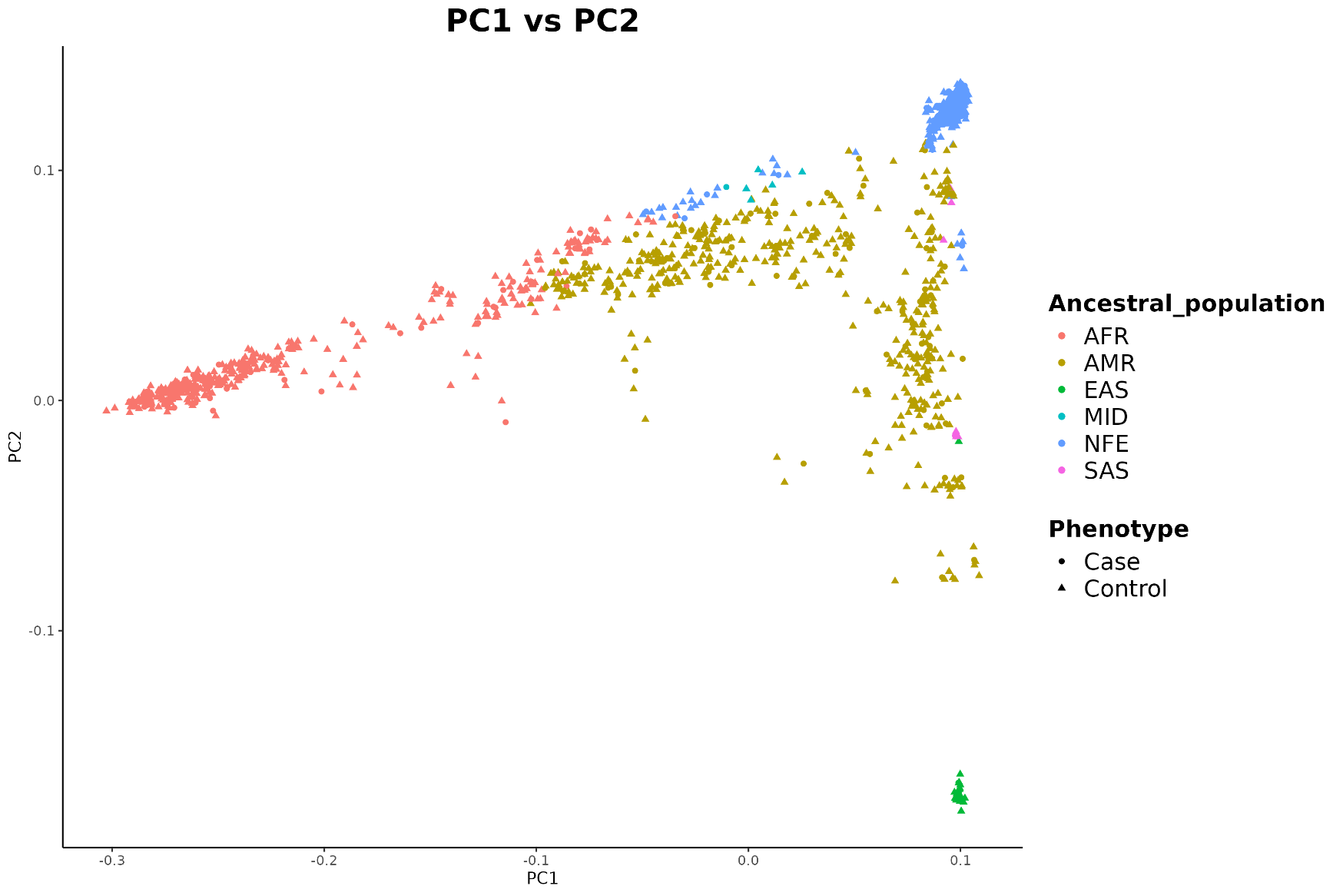

**Extended Data Figure 1. The principal component analysis (PCA) plot of 301 cases and 1,505 matched controls from whole genome sequencing data.** The first principal component (PC1) is on the x-axis and the second principal component (PC2) is on the y-axis. Each point represents an individual, color-coded by predicted population and shaped by phenotype status (case or control). Predicted populations: AFR (African); AMR (Admixed American); EAS (East Asian); NFE (Non-Finnish European); SAS (South Asian).

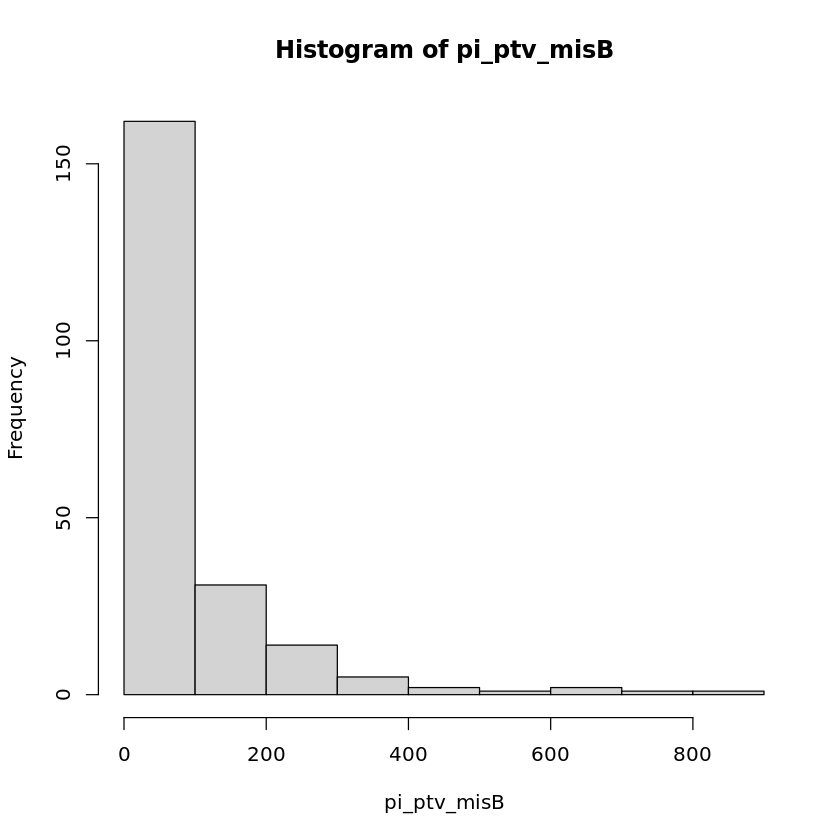

**Extended Data Figure 2. Histogram of the estimated number of risk genes (pi) in 1,000 permutations.** The x-axis represents the estimated number of risk genes calculated based on the number of deleterious variants (pi_ptv_misB): protein-truncating variants (PTVs); missense variants with variants with an MPC (Missense Badness, PolyPhen-2, and Constraint) score ≥ 2 (MisB) , and the y-axis represents the frequency of these estimates across the permutations. The histogram shows the distribution of the estimated number of risk genes.

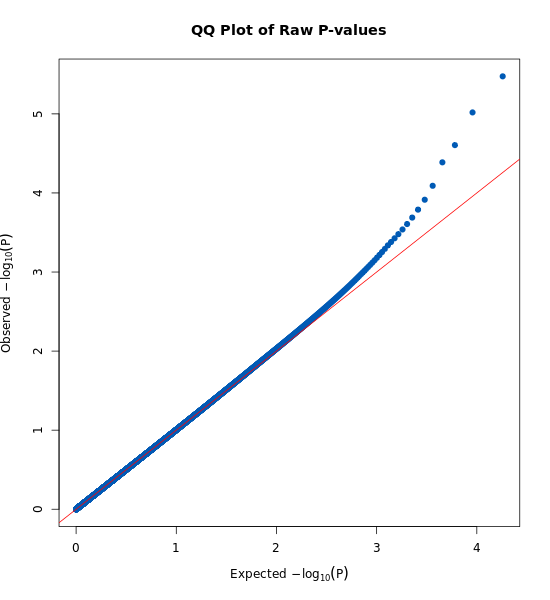

**Extended Data Figure 3. Quantile-quantile plots of TADA (Transmitted and De Novo Association) analysis.** The plot compares the observed -log_10_(*P* values) against the expected -log_10_(*P* values) under the null hypothesis. The red diagonal line represents the expected distribution of *P* values if there is no association.

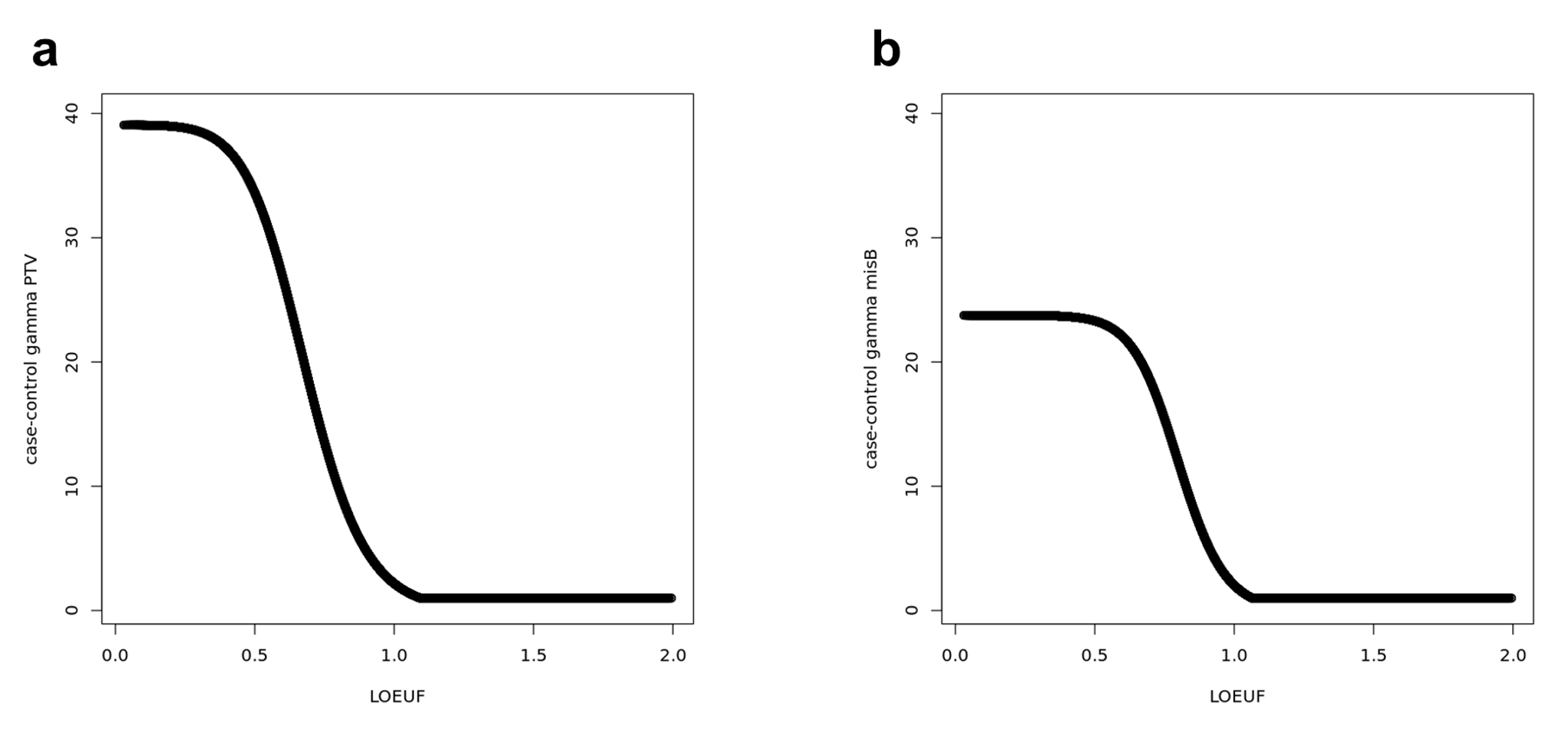

**Extended Data Figure 4. Rolling-average prior risk (gamma) of deleterious variants.** Panel (**a**) shows the case-control gamma PTV (Protein-Truncating Variants) plotted against the LOEUF (Loss-of-function observed/expected upper bound fraction) score. Panel (**b**) shows the case-control gamma MisB, missense variants with variants with an MPC (Missense Badness, PolyPhen-2, and Constraint) score ≥ 2, plotted against the LOEUF score. The y-axis represents the gamma value, and the x-axis represents the LOEUF score.
